# Real-world serologic responses to Extended-interval and Heterologous COVID-19 mRNA vaccination in Frail Elderly - Interim report from a prospective observational cohort study

**DOI:** 10.1101/2021.09.16.21263704

**Authors:** Donald C. Vinh, Jean-Philippe Gouin, Diana Cruz-Santiago, Michelle Canac-Marquis, Stéphane Bernier, Florian Bobeuf, Avik Sengupta, Jean-Philippe Brassard, Alyssa Guerra, Robert Dziarmaga, Anna Perez, Yichun Sun, Yongbiao Li, Lucie Roussel, Mélanie J. Langelier, Danbing Ke, Corey Arnold, Martin Pelchat, Marc-André Langlois, Timothy G. Evans, Xun Zhang, Bruce D. Mazer, on behalf of the COVID-19 Immunity Task Force and UNCoVER Investigators

## Abstract

**Background:** The Coronavirus disease 2019 (Covid-19) pandemic, caused by severe acute respiratory syndrome coronavirus 2 (SARS-CoV-2), has prompted accelerated vaccines development. Their use was prioritized to protect the most vulnerable, notably, the elderly. Because of fluctuations in vaccine availability, strategies such as delayed second dose and heterologous prime-boost have been employed. The effectiveness of these strategies in the frail elderly are unknown.

**Methods:** In this real-world vaccination study, under a government-decreed rationing strategy, elderly adults residing in long-term care facilities, with or without previously-documented SARS-CoV-2 infection, were administered homologous or heterologous mRNA vaccines, with an extended 16-week interval between doses. Clinical data and blood were serially collected during and after this interval period. Sera were tested for SARS-CoV-2-specific IgG antibodies (to trimeric S; RBD; nucleocapsid) by automated chemiluminescent ELISA.

**Findings:** After a significant increase 4 weeks post-prime dose, there was a significant decline in anti-RBD and anti-S IgG levels until the boost dose, followed by an increase 4 weeks later. Previously uninfected individuals exhibited lower antibody responses up to 16 weeks post-prime dose, but achieved comparable levels to previously infected counterparts by 4 weeks post-second dose. Individuals primed with BNT162b2 exhibited larger decrease in anti-RBD and anti-S IgG levels with 16-week interval between doses, compared to those who received mRNA-1273. No differences in antibody levels 4 weeks after the second dose were noted between the two vaccines, in either homologous or heterologous combinations.

**Interpretations:** These interim results of this ongoing longitudinal study show that, among frail elderly, neither age, sex, nor comorbidity affect antigenicity of mRNA-based COVID vaccines, but previous SARS-CoV-2 infection and type of mRNA vaccine influenced antibody responses when used with a 16-week interval between doses. Homologous/heterologous use of mRNA vaccines was not associated with significant differences in antibody responses 4 weeks following second dose, supporting their interchangeability.

**Funding:** This project was supported by funding from the Public Health Agency of Canada, through the Vaccine Surveillance Reference group and the COVID-19 Immunity Task Force (CITF).

## Introduction

The severe acute respiratory syndrome coronavirus 2 (SARS-CoV-2) pandemic has been devastating, particularly for elderly individuals^1, 2^. Accelerated vaccine development via different platforms, including those that are mRNA-based (mRNA-1273 or BNT162b2), are a significant advancement in the fight against the Coronavirus disease 19 (COVID-19) pandemic. Unfortunately, global demand has outweighed supply in many countries, resulting in the common public health strategy to administer the first dose of vaccine to as many at-risk individuals as possible up front^3^; this resulted in delayed administration of the second dose (relative to what was done in clinical trials) and heterologous prime-boost vaccination (whereby the second dose is different from the first dose, or “mixing”). The serologic response under these approaches is unknown, particularly in the frail elderly.

Here, in the UNCoVER (UNderstanding Co-V2 Vaccination in Elderly Residents) study, we evaluated the serological responses during this real-world vaccinating strategy in long term care facility (LTC)-residing elderly and report its interim results.

## Methods

### Study Procedure and Participants

In Québec, Canada, the COVID-19 vaccination campaign was launched by the provincial Ministry of Health on December 14, 2020, with prioritization of available mRNA-1273 or BNT162b2 doses to the elderly residing in LTC. This study was conducted in 12 LTC facilities of the Montréal Centre-Sud – Integrated University Health and Social Services Centre (CIUSSS du Centre-Sud-de-l’Île-de-Montréal) and was approved by its research ethics board (Comité d’éthique de la recherche Vieillissement-Neuroimagerie, protocol number 20-21-36 MP). The LTC facilities involved (termed residential and long-term care centres, CHSLD) focus primarily on the care of elderly people requiring more than 3 hours of care daily. Because of the limited vaccine supply, the province-wide prioritization strategy consisted of administering the first dose to as many LTC elderly as possible, which necessitated a delayed administration of the second dose up to 16 weeks after the first dose, rather than the 3- or 4-weeks interval in the clinical trials. At time of the second dose, the mRNA vaccine used could be either homologous or heterologous to the first one, based on availability. All elderly residents in participating LTC were eligible. About 71% of the residents contacted by the research team agreed to participate in the study. Participating LTC elderly residents or their legally authorized representative provided informed consent before enrollment. Clinical data was collected at baseline and blood samples were collected at the following time points: Prior to 1^st^ dose (t1); at approximately 4 weeks after the 1^st^ dose, which coincides with the timing of the putative second dose, as done in the clinical trials (t2); at 6 to 10 weeks after 1^st^ dose (t3); at the time of actual administration of the 2^nd^ dose 16 weeks after prime dose (t4); and at 4 weeks after 2^nd^ dose (t5); these time measurements constitute this interim report. Longitudinal collection at later time points is in progress.

### Samples

Blood was collected in Acid Citrate Dextrose (ACD) from participants and processed within 6 hours of collection. Plasma was stored at -80°C until ready for batch testing.

### Assessment of SARS-CoV-2 antibody responses

Briefly, longitudinal serologic responses to vaccines were measured by automated chemiluminescent ELISA to detect IgG to SARS-CoV-2 trimeric spike (S), nucleoprotein (N) and the receptor binding domain (RBD) antigens to discriminate between vaccine-induced antibody response and convalescence from natural SARS-CoV-2 infection. Scaled luminescence values were measured, as detailed in Supplementary Material.

### Statistical Analysis

Participants’ demographic and clinical characteristics were reported by proportions for categorical data and by means (ranges) for continuous data. The mean response profiles [for S, R and N] were displayed visually. Linear mixed effects models were used to evaluate change in antibody levels over time, accounting for within-individual variability, and to test interaction between change in antibody levels over time and vaccine homology or previous infection status. Moderation analysis examined the effects of prior SARS-CoV-2 infection, vaccine type, vaccine homology, age, sex, and medical comorbidities.

## Results

### LTC Population

This real-world observational study was conducted on two cohorts: In the first cohort (termed T1+), blood samples were collected prior to the administration of the first vaccine dose and longitudinally thereafter. In the second cohort (T2+), blood samples were collected starting at the t2 time point; because of this difference in sampling, the T2+ cohort was used to confirm findings observed in T1+, when applicable. A total of 228 subjects were enrolled between December 30, 2020 and February 16, 2021. Fourteen subjects were excluded because they did not receive the second vaccine dose, due to either death, refusal of the second vaccine, or withdrawal from the study. An additional 29 subjects were excluded because they were less than 65 years old (i.e. not elderly). Thus, the total study sample consists of 185 elderly persons: Among these participants, the median age was 83 years, 69·2% were female, 89·7% were White, 46·5% had been previously diagnosed with SARS-CoV-2 infection (confirmed by RT-PCR in the respective diagnostic laboratory), and 98% had at least one coexisting condition. Based on the Clinical Frailty Scale^4^, the mean score of the included participants was 6·57 (median: 7, range: 1 to 8). The first vaccine dose was mRNA-1273 in 80·5% of subjects and BNT162b2 in 19·5%. The second dose was mRNA-1273 in 35·1% and BNT162b2 in 64·9% of subjects. Comparison of sub-groups demonstrated that T1+ (n=78) and T2+ (n=107) were comparable in age, sex, race, comorbidity, and frailty, although differences were observed in proportion with previous infection and vaccine homology (Table 1).

**Table 1:**
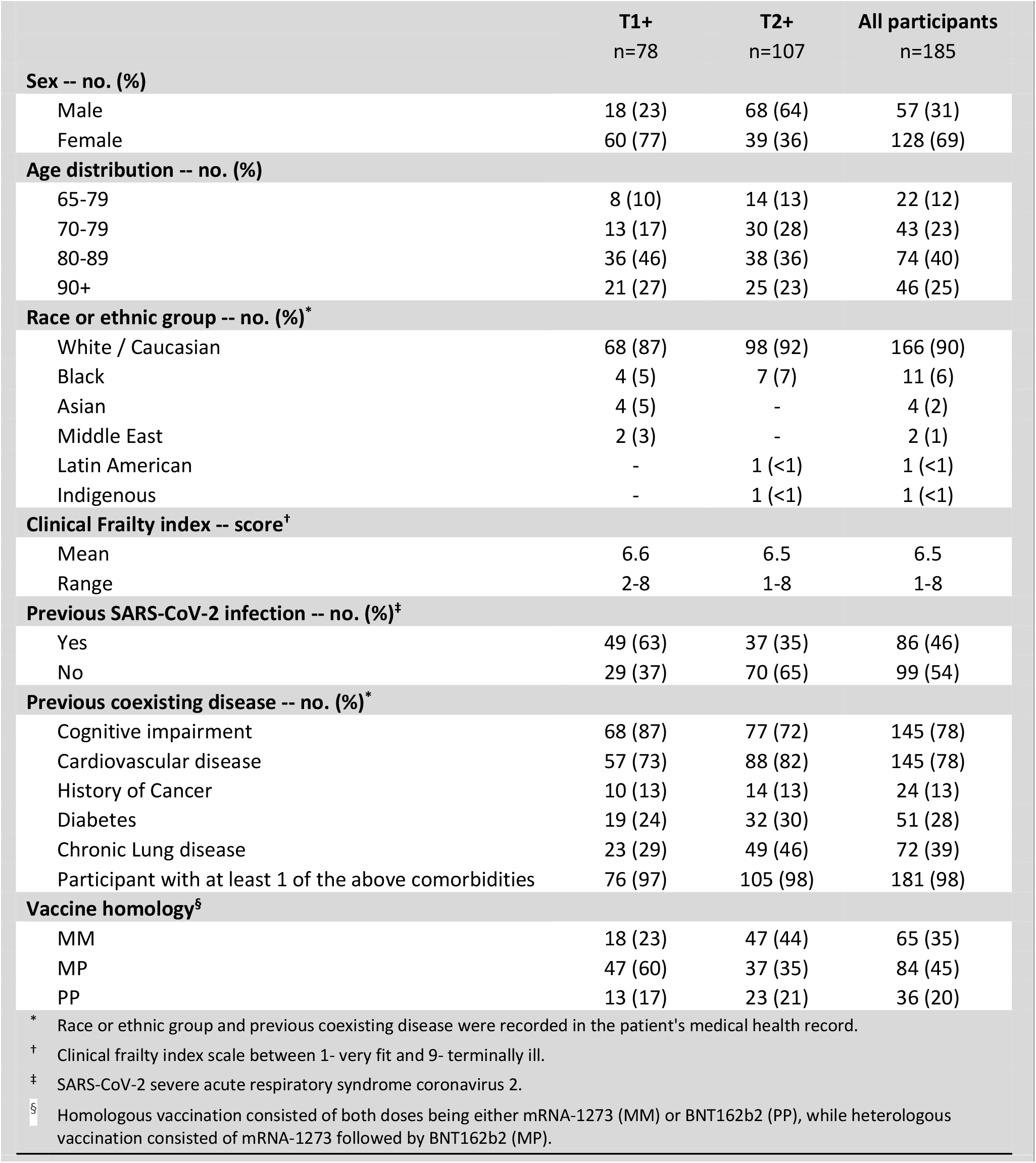
Characteristics of the Participants at Baseline.

|  | T1+<br>n=78 | T2+<br>n=107 | All participants<br>n=185 |
| --- | --- | --- | --- |
| **Sex -- no. (%)** |  |  |  |
| Male | 18 (23) | 68 (64) | 57 (31) |
| Female | 60 (77) | 39 (36) | 128 (69) |
| **Age distribution -- no. (%)** |  |  |  |
| 65-79 | 8 (10) | 14 (13) | 22 (12) |
| 70-79 | 13 (17) | 30 (28) | 43 (23) |
| 80-89 | 36 (46) | 38 (36) | 74 (40) |
| 90+ | 21 (27) | 25 (23) | 46 (25) |
| **Race or ethnic group -- no. (%)<sup>\*</sup>** |  |  |  |
| White / Caucasian | 68 (87) | 98 (92) | 166 (90) |
| Black | 4 (5) | 7 (7) | 11 (6) |
| Asian | 4 (5) | - | 4 (2) |
| Middle East | 2 (3) | - | 2 (1) |
| Latin American | - | 1 (<1) | 1 (<1) |
| Indigenous | - | 1 (<1) | 1 (<1) |
| **Clinical Frailty index -- score<sup>†</sup>** |  |  |  |
| Mean | 6.6 | 6.5 | 6.5 |
| Range | 2-8 | 1-8 | 1-8 |
| **Previous SARS-CoV-2 infection -- no. (%)<sup>‡</sup>** |  |  |  |
| Yes | 49 (63) | 37 (35) | 86 (46) |
| No | 29 (37) | 70 (65) | 99 (54) |
| **Previous coexisting disease -- no. (%)<sup>\*</sup>** |  |  |  |
| Cognitive impairment | 68 (87) | 77 (72) | 145 (78) |
| Cardiovascular disease | 57 (73) | 88 (82) | 145 (78) |
| History of Cancer | 10 (13) | 14 (13) | 24 (13) |
| Diabetes | 19 (24) | 32 (30) | 51 (28) |
| Chronic Lung disease | 23 (29) | 49 (46) | 72 (39) |
| Participant with at least 1 of the above comorbidities | 76 (97) | 105 (98) | 181 (98) |
| **Vaccine homology<sup>§</sup>** |  |  |  |
| MM | 18 (23) | 47 (44) | 65 (35) |
| MP | 47 (60) | 37 (35) | 84 (45) |
| PP | 13 (17) | 23 (21) | 36 (20) |

\* Race or ethnic group and previous coexisting disease were recorded in the patient's medical health record.
† Clinical frailty index scale between 1- very fit and 9- terminally ill.
‡ SARS-CoV-2 severe acute respiratory syndrome coronavirus 2.
§ Homologous vaccination consisted of both doses being either mRNA-1273 (MM) or BNT162b2 (PP), while heterologous vaccination consisted of mRNA-1273 followed by BNT162b2 (MP).

### Longitudinal serologic responses based on previous infection status

In the T1+ group, 49 (62·8%) of participants had previous, microbiologically-confirmed natural infection with SARS-CoV-2. These statuses were reflected in the mean anti-N IgG levels at baseline (t1): 0·49 (uninfected) and 1·35 (infected) [p<0·001] [Figure 1]. Those who were previously infected maintained anti-N reactivity over the subsequent time points and at higher levels than those previously uninfected. For the anti-S IgG response, contrasting mean levels were noted at t1 based on status of previous infection: 0·48 (uninfected); 1·78 (infected) [p<0·001]. However, comparable levels were detected at t2 (4 weeks after first dose) and t3 (6-10 weeks after first dose). Notably, at t4 (16 weeks after first dose), the mean anti-S IgG response in those previously uninfected declined to lower levels than those who had recovered from natural infection (1·15 vs. 1·56, respectively [p<0·001]). This difference resolved by t5 (4 weeks after second dose). anti-RBD IgG levels at baseline (t1) were significantly higher in the previously infected subjects: 0·32 (uninfected) and 1·23 (infected) [p<0·001]. Those previously infected maintained statistically significantly higher mean anti-RBD IgG activity over the following 16 weeks (t2 to t4), compared to those previously uninfected, in whom levels declined at t4 (16 weeks). However, by 4 weeks after boost immunization (t5), the anti-RBD IgG levels were comparable between the two groups. The dynamics of the anti-N, anti-S, and anti-RBD IgG levels, starting at 4 weeks following the first dose, were confirmed in the T2+ cohort (Supplemental Figure 1), including the decline in the latter two responses at t4.

**Figure 1.**
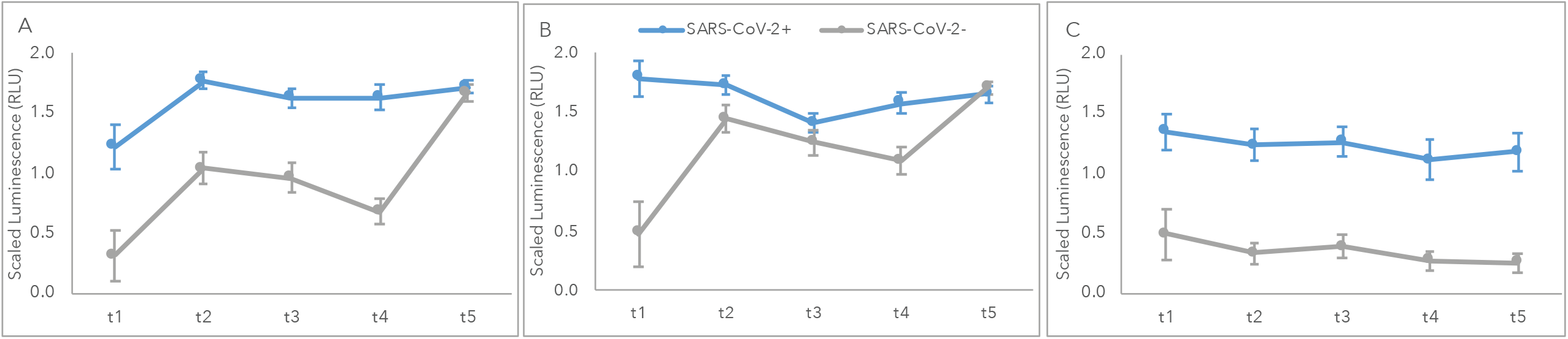
Antibody responses based on previous infection with SARS-CoV-2. Antibody responses to (A) RBD, (B) S and (C) N antigens, based on previous infection with SARS-CoV-2. RLU: relative light units.

### Antigenicity of Homologous vs. Heterologous vaccination approaches

Homologous vaccination consisted of both doses being either mRNA-1273 (MM) or BNT162b2 (PP), while heterologous vaccination consisted of mRNA-1273 followed by BNT162b2 (MP). There were no participants who received BNT162b2 followed by mRNA-1273 in the current study. In the T1+ cohort, the proportion of subjects receiving MM, PP, or MP were 23·1%, 16·7%, or 60·3%, respectively. At t2 (4 weeks after first dose), the mean anti-RBD IgG increased from 0·74 to 1·44 in the MM group, from 0·90 to 1·46 in the PP group, and from 0·94 to 1·60 in the MP group [Figure 2]. The mean anti-RBD IgG reactivity over the ensuing 6 to 10 weeks (t3) was 1·44 (MM), 1·39 (PP), and 1·45 (MP). At the time of the second dose (t4, i.e., 16 weeks after the first dose), those immunized with PP had significantly lower anti-RBD IgG levels, compared to persons receiving MM or MP (0·63 vs. 1·32 or 1·42, respectively; p=0·002). This difference was similarly seen in anti-S IgG responses (1·02 vs. 1·45 or 1·49; p=0·012). Mean levels of anti-N IgG were not significantly different across time points. At 4 weeks after boost immunization (t5), anti-RBD IgG responses in those receiving BNT162b2 were similar to those who had received mRNA-1273. Similar IgG responses were observed for the T2+ cohort, and the total group combining T1+ and T2+ [Supplemental Figure 2]. There was thus no difference between heterologous and homologous mRNA vaccination 4 weeks post booster dose.

**Figure 2.**
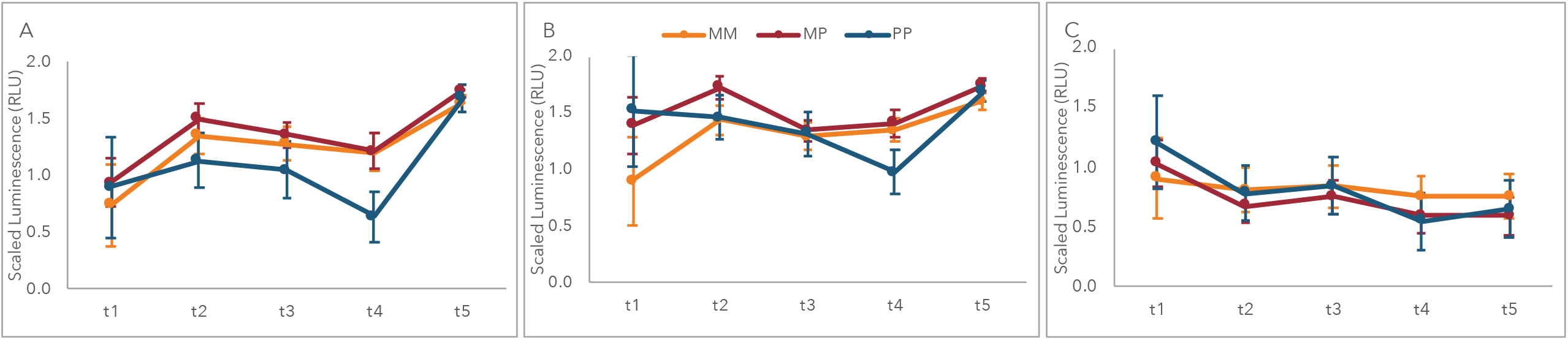
Antibody responses based on homologous vs. heterologous use of mRNA vaccines. Antibody responses to (A) RBD, (B) S and (C) N antigens, based on homologous vs. heterologous use of mRNA vaccines. M: Moderna (mR-NA-1273). P: Pfizer (BNT162b2). RLU: relative light units.

To determine if previous infection status impacted the antigenicity of these vaccine strategies, we analyzed separately the impact of vaccine homology in those without and with a history of SARS-CoV-2 infection [Figure 3]. As expected, the anti-N IgG levels were higher in the latter group throughout the time period. In recovered patients, anti-RBD IgG and anti-S levels were consistently higher until t5 (4 weeks after second dose), compared to uninfected patients. Among participants who received BNT162b2, both previously infected and uninfected individuals experience a decrease in anti-RBD IgG from t2-t4. In contrast, among those who receive mRNA-1273, individuals with prior infection showed no significant change in antibody responses from t2-t4, while uninfected individuals experience a decrease in anti-RBD IgG, albeit less than their counterparts who received BNT162b2. At 4 weeks post second dose, there was an increase in antibody levels in all groups; no differences related to vaccine type, vaccine homology, and prior SARS-CoV-2 infection were observed.

**Figure 3.**
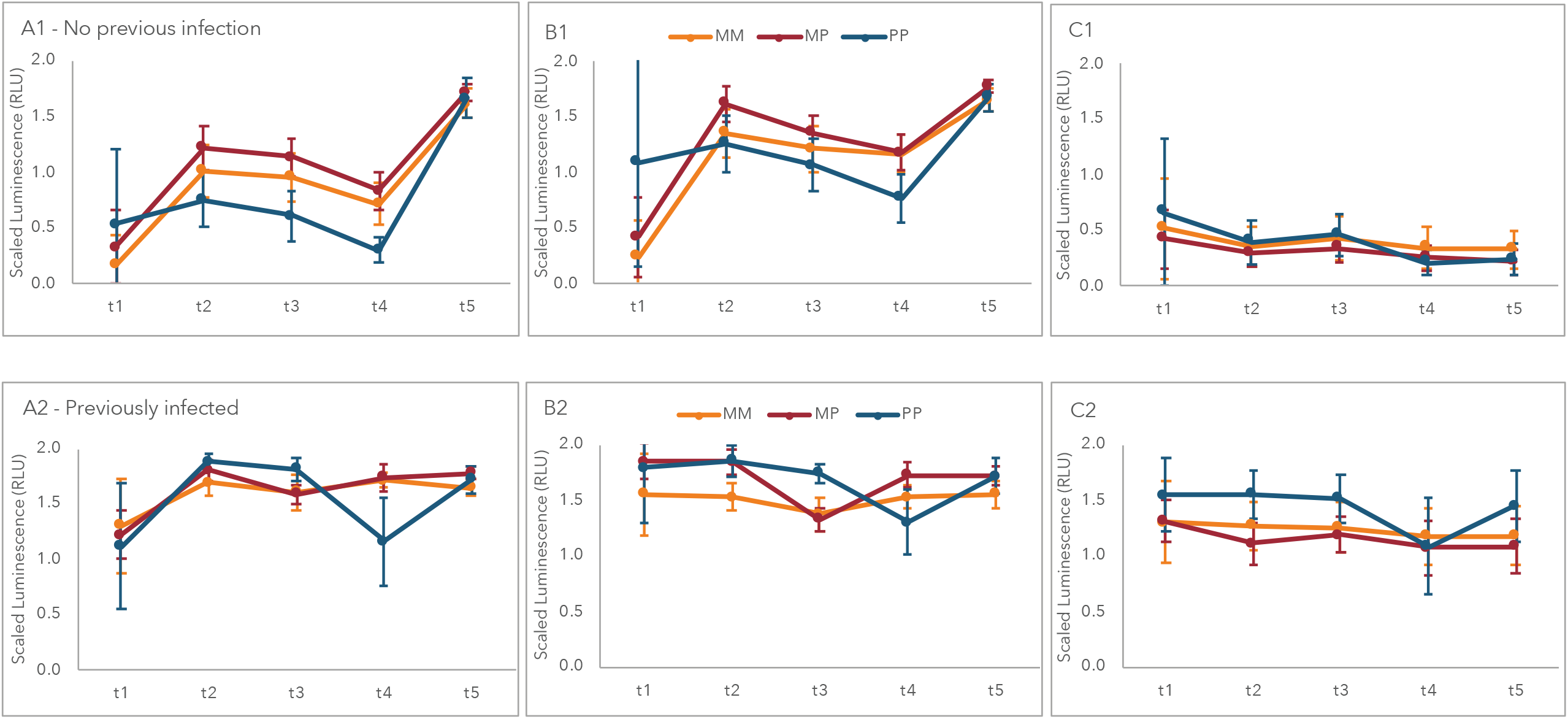
Antibody responses to homologous vs. heterologous use of mRNA vaccines based on previous infection status. Antibody responses to (A1, A2) RBD, (B1, B2) S and (C1, C2) N antigens, with homologous vs. heterologous use of mRNA vaccines, based on absence (top row) or presence (bottom row) of previous SARS-CoV2 infection status. M: Moderna (mRNA-1273). P: Pfizer (BNT162b2). RLU: relative light units.

### Impact of age, sex, or comorbidity on antibody responses

To determine whether antigenicity varies with age, we categorized T1+ subjects’ age into decades and assessed their serologic response. The mean anti-N, anti-S, and anti-RBD IgG levels were comparable, at all time points, across the age categories [Supplemental Figure 3a]. These findings were confirmed in the T2+ cohort. Similarly, no differences in responses were observed when stratifying by sex or by comorbidity in both cohorts [Supplemental Figures 3b-3c]. Comparable antibody responses were observed between age groups or sex when stratifying based on previous infection status or by homologous/heterologous use of vaccine [Supplemental Figure 4-7].

## Discussion

During the first waves of the pandemic, the elderly population was at risk for life-threatening COVID-19 disease. Because the mRNA-based vaccines (mRNA-1273 (Moderna) and BNT162b2 (Pfizer/BioNTech)) demonstrated significant serologic responses after the first dose in clinical trials^5,^ _6_, coupled with the scarcity of supply globally, public health measures in several countries rationed their vaccines by deferring second doses until high-risk demographics received their first dose. This approach resulted in extended intervals of 16 weeks between doses in Quebec, in contrast to the 3-4 weeks interval studied in clinical trials. While this policy expanded protection widely, resulting in decreased hospitalizations and deaths^7^, the antigenicity of this strategy in prioritized, vaccinated LTC elderly recipients is unknown. As well, because of the variability in supply, the boost doses administered were either the same (homologous) or different (heterologous) from the prime doses, although data on the inter-changeability of the mRNA vaccines is lacking. Lastly, because of the urgency of vaccination of LTC elderly, all residents wishing to be vaccinated received it, regardless of their previous SARS-CoV-2 infection status. How previous infection impacts vaccine response in the frail elderly is unknown. This study allowed us to evaluate the real-world effectiveness of these public health decisions, using a discovery cohort (T1+), with validation in a confirmatory cohort (T2+).

We show that among the frail institutionalized elderly, there is no age-, sex-nor comorbidity-based differences in IgG humoral immune responses induced by the mRNA-based SARS-CoV-2 vaccines. Residents of LTC, who are often older and frail, may have vaccine-induced immune responses that are less robust compared to younger individuals due to immunosenescence, a multifactorial process that results in declining immunity with advancing age^8^. Indeed, elderly vaccinees have demonstrated diminished serological responses to COVID-19 vaccination compared to non-elderly ones^9-12^. Here, our analyses demonstrate no variances to the mRNA-based vaccines among the different age decades within these cohorts of frail elderly. Similarly, sex-based differences in antibody responses to vaccines occur in older-aged individuals to certain vaccines (e.g. influenza, zoster, pneumococcus)^13^. No differential responses were noted in our two cohorts. These findings demonstrate that the antigenicity of this vaccination strategy was equally efficacious, regardless of age or sex, at least during the first five months from the first dose.

Previous natural infection with SARS-CoV-2, documented by microbiological testing, occurred in 46.5% of our total participants; this classification was confirmed by anti-N antibody positivity at baseline, with concomitantly higher levels of anti-S and anti-RBD IgG levels. Here, several findings are noteworthy: Firstly, in those with confirmed, previous infection, a two-dose mRNA-based vaccine strategy, separated by 16 weeks between doses, resulted in elevated levels of all SARS-CoV-2-specific IgG responses, at least during the five-month time frame of this report. Secondly, among those who were not previously infected, the anti-RBD IgG responses achieved were significantly lower at all time points prior to the second dose, compared to their counterparts with prior SARS-CoV-2 infection. This effect has been previously noted in the elderly, although in smaller studies^11, 12^. Thirdly, among this infection-naïve group, both anti-S and anti-RBD IgG levels decreased significantly at four months after the first dose. However, administration of the second dose at this time augmented the corresponding antibodies to levels comparable to those with previous infection. This decline was observed in our T1+ cohort and confirmed in our T2+ cohort, and suggests that the extended interval of 16 weeks between doses may be the maximum period permitted to limit waning of these responses, at least in those who had not been previously infected.

In comparing antibody responses under different vaccine mixing strategies, a consistent pattern emerges: use of BNT162b2 as first dose results in a significantly faster decline in anti-S and anti-RBD IgG responses during the 16-week interval between the two vaccine doses, relative to mRNA-1273. Administering BNT162b2 as second dose at this time point, in either homologous (PP) or heterologous (MP) manner, achieves comparable antibody responses to homologous mRNA-1273 use (MM) by 1 month later. A difference in antigenicity between these mRNA vaccines is emerging^14-16^; our study highlights this phenomenon also occurs in the frail elderly, at least following the prime dose. While studies on heterologous vaccine platforms have focused on the immunogenicity of adenoviral-based vaccines with mRNA-based ones^17, 18^, the interchangeability of the mRNA-based vaccines and their heterologous use in the frail elderly have not, to our knowledge, been evaluated in a real-world setting. Our data indicate that there are indeed differences in the kinetics of induced antibody responses between mRNA-1273 and BNT162b2, detectable at four months from first dose. Moreover, homologous use of mRNA-based vaccines is not necessarily equivalent, as shown by the dynamic difference in antibody levels between MM and PP. While our data suggest that the specific heterologous use of mRNA-1273 then BNT162b2 (MP) are comparable to MM, the converse order (BNT162b2 then mRNA-1273, PM) was not used in this study and cannot be compared.

One limitation of this study is the focus on IgG responses. Although immune correlates of protection against COVID-19 are not currently established, the spike protein-specific antibody responses (here, assessed by IgG to trimeric S and RBD antigens) generally correlate with the level of neutralizing antibody activity, which is thought to be a critical determinant of vaccine efficacy. Although the clinical relevance of induced, circulating IgA and IgM responses is less clear, we are currently evaluating these levels. As well, our study does not provide data on clinical effectiveness against infection or disease during the study period. Lastly, cellular immune responses were not included in this interim report, given the potential impact of our findings for nascent vaccination strategies in other countries, although those immunologic studies are ongoing.

In frail elderly individuals, the use of mRNA-based vaccines against COVID-19 can be used with an extended interval of up to 16 weeks between doses. Neither age, sex, or comorbidity appears to impact these serological responses. Robust antibody levels are elicited with either mRNA-1273 or BNT162b2 as first dose, particularly in those with previously documented infection, although the latter vaccine is distinctly associated with a decline in antibody levels in the ensuing 16 weeks prior to the second dose. Under this dosing strategy, comparable antibody levels are achieved with either homologous or heterologous use of these vaccines by one month after the second dose. These data can be used to shape vaccination policies globally, especially in light of vaccine supply shortages.

## Data Availability

De-identified clinical data for the patients in this study might be made available to other investigators after approval by the institutional review board. Requests should be directed to the corresponding author.

## Contributors

DCV: Conceptualisation; data curation; formal analysis; funding acquisition; investigation; methodology; project administration; supervision; validation; visualization; writing-original draft; writing-review & editing.

JPG : Conceptualisation; data curation; formal analysis; funding acquisition; investigation; methodology; project administration; supervision; validation; visualization; writing-original draft; writing-review & editing.

DCS : Conceptualisation; formal analysis; funding acquisition; investigation; project administration; writing-original draft; writing-review & editing.

MCM : Conceptualisation; data curation; formal analysis; funding acquisition; investigation; methodology; project administration; software; supervision; visualization; writing-original draft; writing-review & editing.

SB : investigation; writing-review & editing.

FB : investigation; validation; writing-review & editing.

AS: investigation; validation; writing-review & editing.

JPB: investigation; validation; writing-review & editing.

AG: investigation; writing-review & editing.

RD: investigation; writing-review & editing.

AP: investigation; writing-review & editing.

YS: investigation; writing-review & editing.

YL: investigation; writing-review & editing.

LR: investigation; writing-review & editing.

MJL: investigation; validation; writing-review & editing.

DK: formal analysis; methodology; resources; writing-review & editing.

CA: data curation; formal analysis; methodology; resources; software; writing-review & editing.

MP: data curation; formal analysis; methodology; resources; software; visualization; writing-review & editing.

MAL: data curation; formal analysis; methodology; resources; software; writing-review & editing.

TGE: funding acquisition; writing-review & editing.

XZ: data curation; formal analysis; methodology; resources; software; visualization; writing-original draft; writing-review & editing.

BDM: Conceptualisation; funding acquisition; writing-review & editing.

DCV, JPG, DCS, and MCM accessed and verified the data and had final responsibility for the decision to submit for publication.

## Acknowledgements

We would like to acknowledge the ongoing collaboration of the Centre intégré universitaire de santé et de services sociaux du Centre-Sud-de-l’Île-de-Montréal (CCSMTL) for this study. We would like to thank the CHSLD residents and their family members for their participation.

This project was supported by funding from the Public Health Agency of Canada through the COVID-19 Immunity Task Force (CITF) and by a COVID-19 Rapid Response grant from the Canadian Institute of Health Research (CIHR; #VR2 - 172722) and by a grant supplement by the CITF to M-A Langlois. Production of COVID-19 reagents was financially supported by the NRC’s Pandemic Response Challenge Program.

