## Supplemental Figure 1 for "Real-world serologic responses to Extended-interval and Heterologous COVID-19 mRNA vaccination in Frail Elderly - Interim report from a prospective observational cohort study"

### Supplementary Material

### Methods:

#### Chemiluminescent automated ELISA

Automated chemiluminescent ELISAs were based upon and optimized from assays first described here (10.1126/sciimmunol.abe5511). Hamilton MicroLab Star robotic liquid handlers and plate washer (Biotek 405 TS/LS LHC2, all washes were performed 4x in 100 µl PBS + 0·1% Tween 20 (PBST)) were used for all steps at the University of Ottawa, Faculty of Medicine. All incubation times on the platform were done at room temperature shaking at 500-700 rpm. COVID-19 antigens (Spike trimer, RBD and N), as well as the anti-hIgG#5-HRP fusion antibody were generously provided by Dr. Yves Durocher, National Research Council of Canada (NRC), Montreal. Antigen production was described here (10.1126/sciimmunol.abe5511). Antigens (spike trimer, RBD, N) were diluted in Phosphate Buffered Saline (PBS) and dispensed into wells of a 384-well high-binding polystyrene Nunc plate (ThermoFisher, #460372) at a final amount of 50 ng/well. The plates were centrifuged in a plate spinner for 1 minute to ensure even coating, incubated overnight rocking at 4°C and washed. Wells were blocked with 80 µl of 3% w/v skim milk powder dissolved in PBST for one hour and then washed. Samples were diluted at 1:100 (for Spike, RBD and NP analysis) and 1:10000 (for analysis of Spike and RBD in vaccinated individuals) in 1% w/v skim milk powder dissolved in PBST and 10 µL was added to each well from a 96-well source plate. Standard curves and controls were diluted as per Supplemental Table 1 in 1% w/v skim milk powder dissolved in PBST and 10 µL was added to each plate. Plates were incubated for 2 hours and wells were washed with PBST. Secondary antibodies (anti-human IgG-HRP (NRC anti-hIgG#5-HRP fusion), anti-human IgA-HRP (Jackson ImmunoResearch, 109-035-011), anti-human IgM-HRP (Jackson ImmunoResearch, 109-035-129)) were diluted at 1:5400, 1:8000 and 1:9600, respectively, in 1% w/v skim milk powder in PBST and 10 µl was added to each well. After incubation for 1 hour, the wells were washed and 10 µl of ELISA Pico substrate (ThermoFisher Scientific, #37069, diluted 1:2 in MilliQ $H_2O$) was dispensed to each well. After a 5 min incubation, plates were read on an NEO2 (Bio-Tek) plate reader at 20 ms/well at a read height of 1·0 mm.

#### Data Processing

For consistency in data processing, a constant plate layout containing all of the controls and standard curves in quadruplicates was established. Luminescence values obtained from the antigen-specific standard curve were modelized using a four-parameter log-logistic function to identify the inflection point. Blank-subtracted luminescence values for samples were scaled in relation to the luminescence value corresponding to the inflection point to allow data normalization for subsequent processing. All data processing was performed in R. Density distributions were determined using defaults density function in R with default parameters. Standard curve processing was determined using "LL.4" four-parameter log-logistic self-starter function from the "drc" package in R used with default parameters. Density distributions were performed with the-density function in R with default parameters.

**Supplemental Table 1: Standard curves and controls for automated chemiluminescent ELISA:**

| | | | Concentrations (ng/µL) | | | | | | | |
|---|---|---|---|---|---|---|---|---|---|---|
| | **Reagent** | **Source** | STD 1 | STD2 | STD3 | STD4 | STD5 | STD6 | STD7 | STD8 |
| Standard Curves | anti-Spike, anti-RBD | Absolute Antibody CR3022 (Ab01680-10.0) | 0·5 | 0·25 | 0·125 | 0·0625 | 0·03125 | 0·015625 | 0·00390625 | 0·000976563 |
| | anti-NP | GenScript HC2003 (A02039) | 0·5 | 0·25 | 0·125 | 0·0625 | 0·03125 | 0·015625 | 0·00390625 | 0·000976563 |

| | | | Concentrations | | |
|---|---|---|---|---|---|
| Controls | Pooled Positive Serum | In House | 1:50 | 1:100 | 1:250 |
| | Pooled Negative Serum | in House | 1:50 | 1:100 | |
| | IgG from Human Serum | Sigma (I4506) | 1.0 µg/mL | | |

### Supplementary Figure Legend:

Supplementary Figure 1. Antibody responses based on previous infection with SARS-CoV-2 in the confirmatory (T2+) cohort
Supplementary Figure 2. Antibody responses based on homologous vs. heterologous use of mRNA vaccines in the confirmatory (T2+) cohort
Supplementary Figure 3. Antibody responses based on age, sex and comorbidity
Supplementary Figure 4. Antibody responses based on age stratified by previous infection status
Supplementary Figure 5. Antibody responses based on age stratified by homologous vs. heterologous use of mRNA vaccines
Supplementary Figure 6. Antibody responses based on sex stratified by previous infection status
Supplementary Figure 7. Antibody responses based on sex stratified by homologous vs. heterologous use of mRNA vaccines

**Supplemental Figure 1.** Antibody responses based on previous infection with SARS-CoV-2 in the confirmatory (T2+) cohort

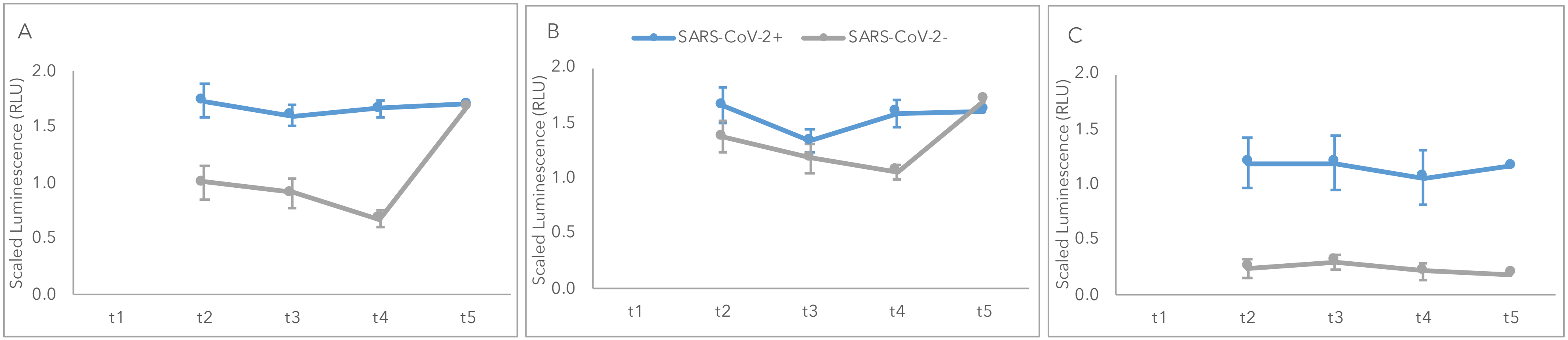

Antibody responses to (A) RBD, (B) S and (C) N antigens, based on previous infection with SARS-CoV-2. RLU: relative light units.

**Supplemental Figure 2.** Antibody responses based on homologous vs. heterologous use of mRNA vaccines in the confirmatory (T2+) cohort

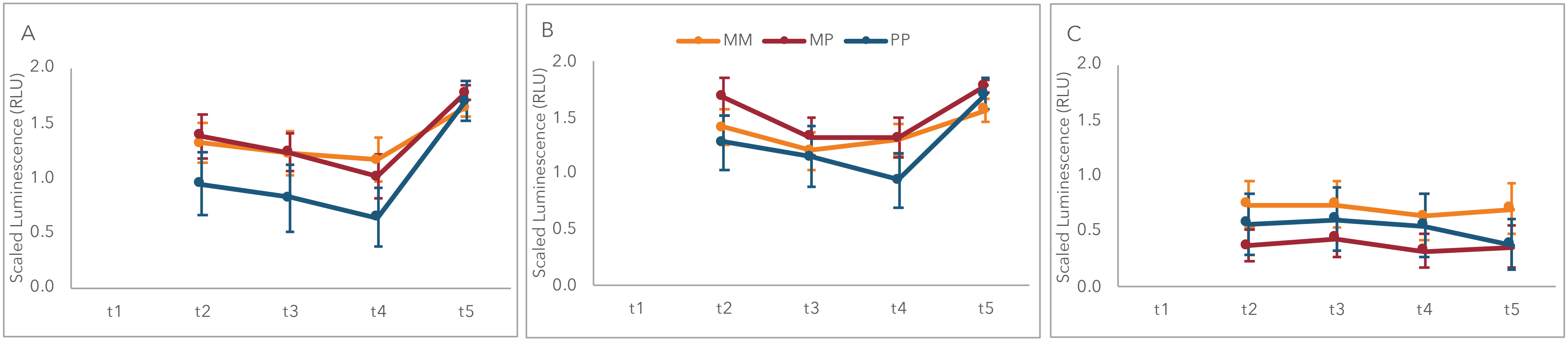

Antibody responses to (A) RBD, (B) S and (C) N antigens, based on homologous vs. heterologous use of mRNA vaccines. M: Moderna (mRNA-1273). P: Pfizer (BNT162b2). RLU: relative light units.

**Supplemental Figure 3a.** Antibody responses based on age

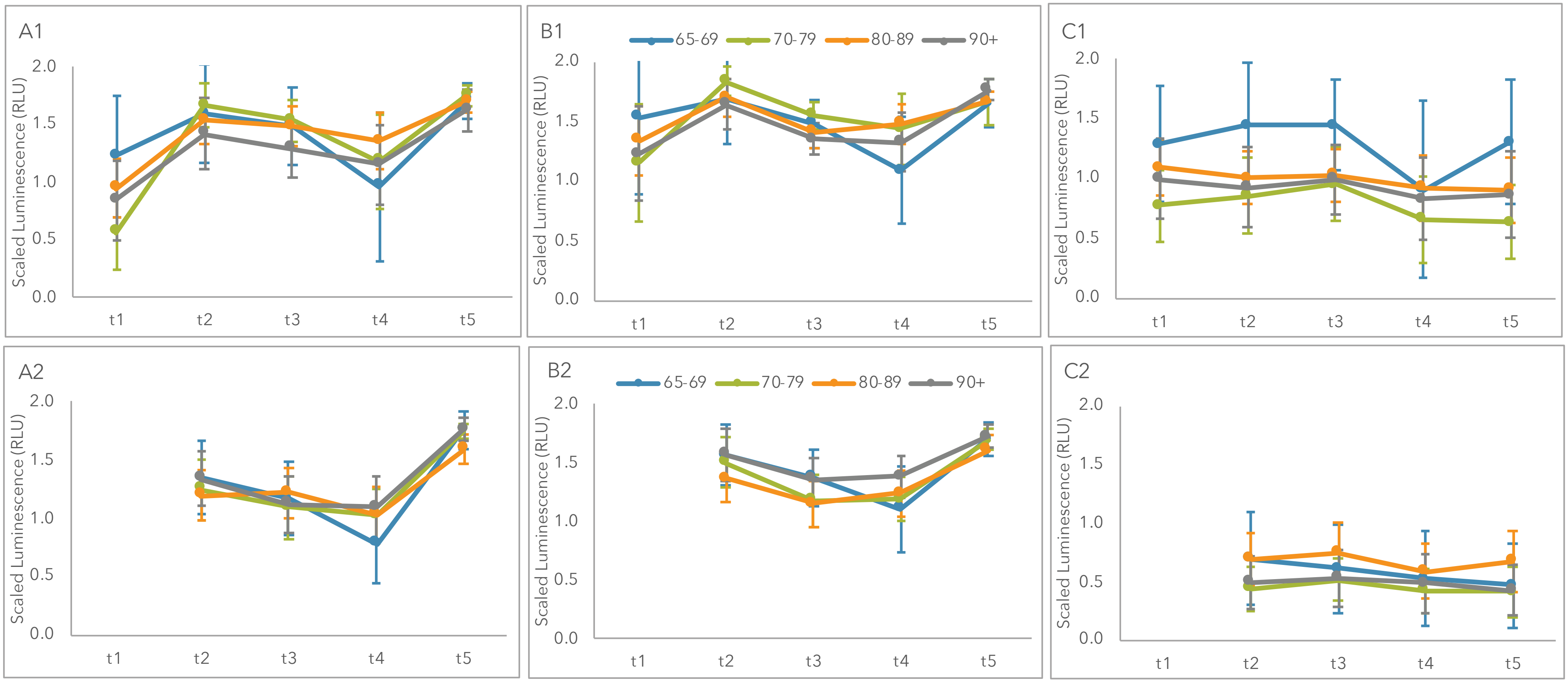

Antibody responses to (A1, A2) RBD, (B1, B2) S and (C1, C2) N antigens, based on age group in T1+ (A1, B1, C1) and T2+ (A2, B2, C2) cohorts. RLU: relative light units.

**Supplemental Figure 3b.** Antibody responses based on sex

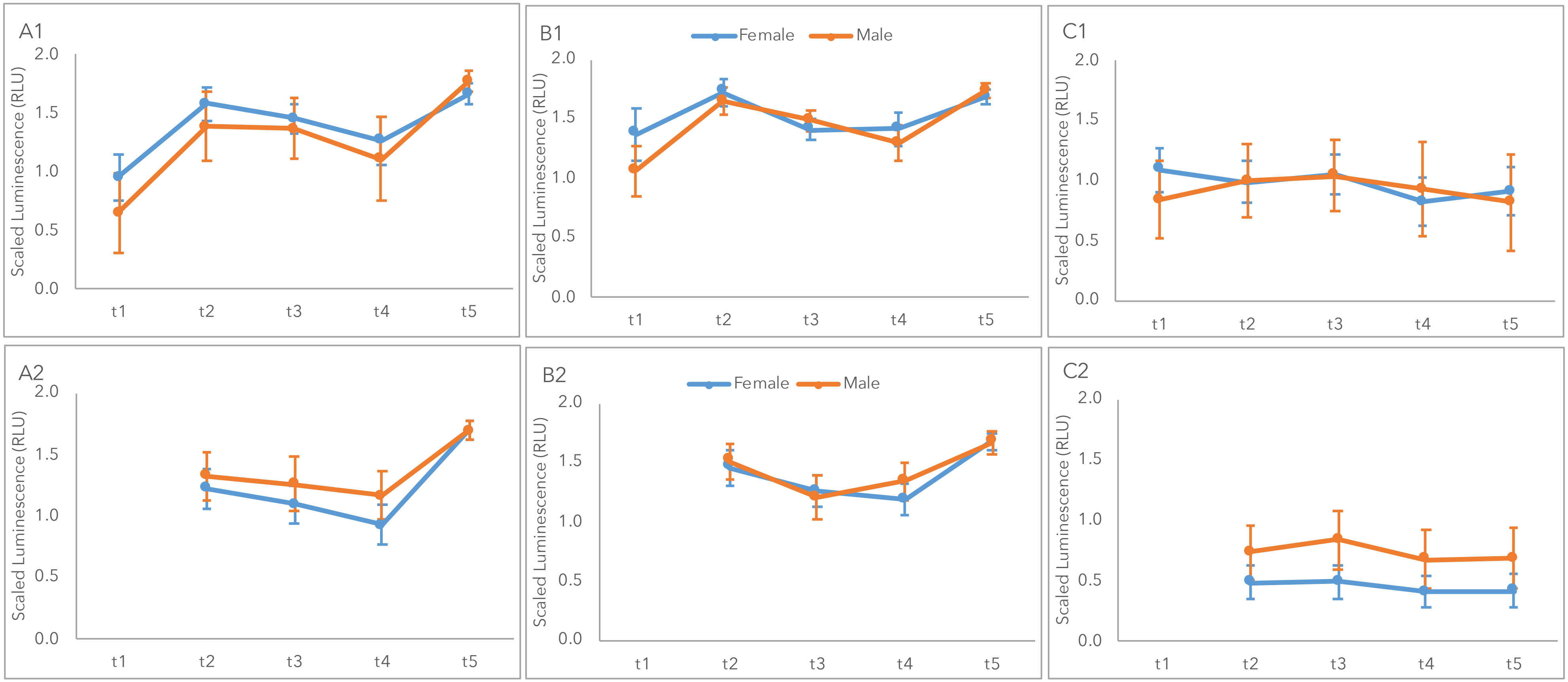

Antibody responses to (A1, A2) RBD, (B1, B2) S and (C1, C2) N antigens, based on sex in T1+ (A1, B1, C1) and T2+ (A2, B2, C2) cohorts. RLU: relative light units.

**Supplemental Figure 3c.** Antibody responses based on comorbidity

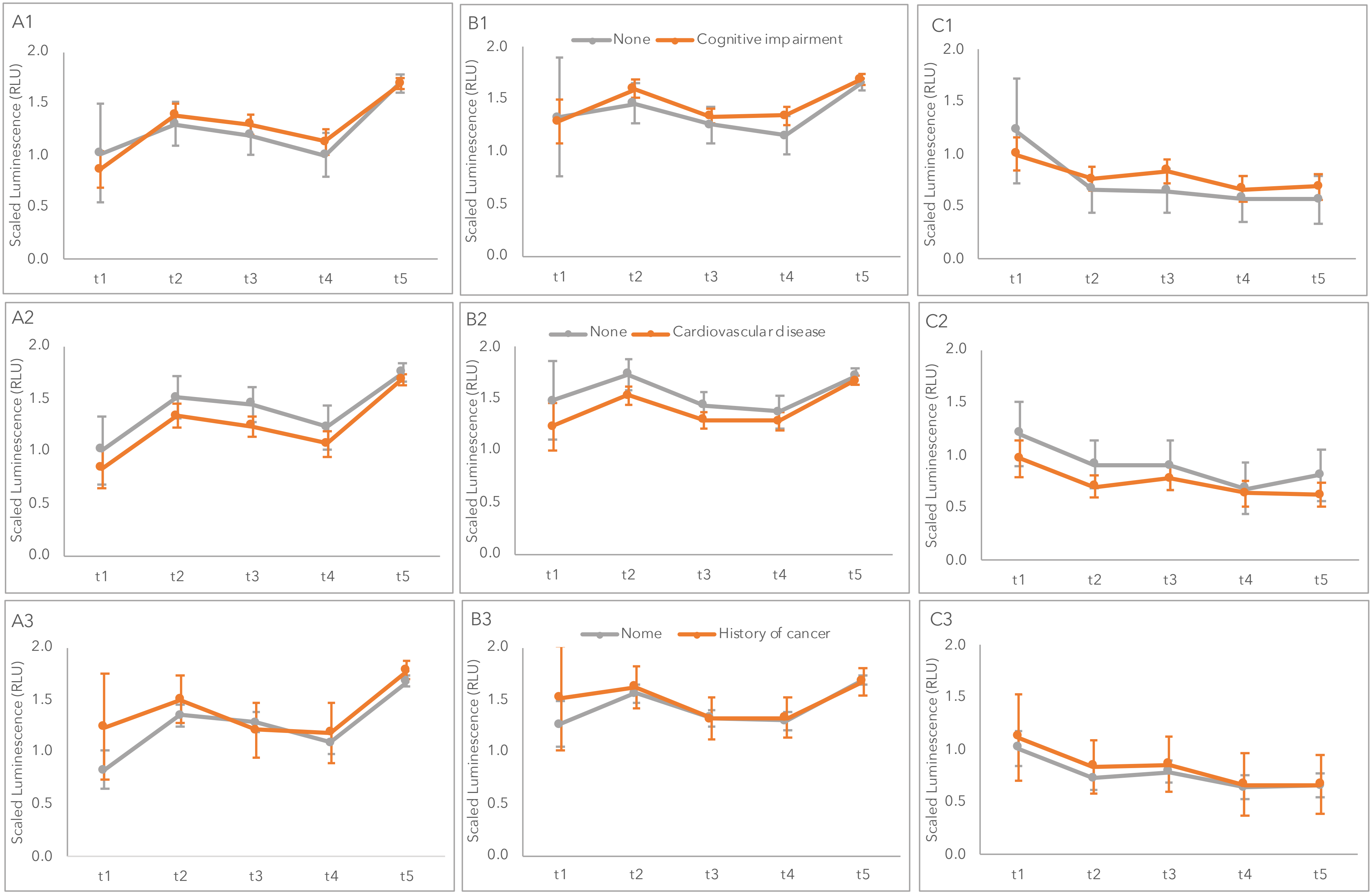

**Supplemental Figure 3c.** Antibody responses based on comorbidity (cont'd)

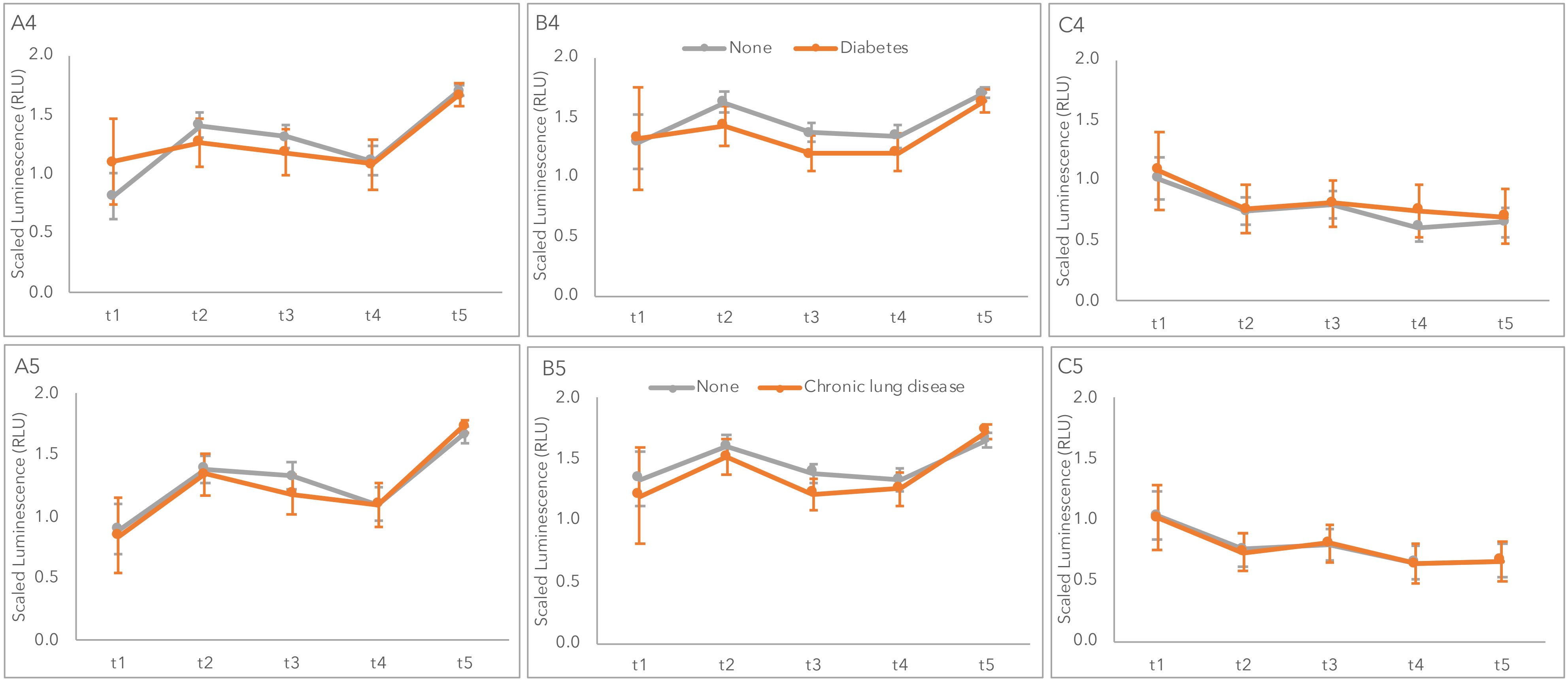

Antibody responses to (A1, A2, A3, A4, A5) RBD, (B1, B2, B3, B4, B5) S and (C1, C2, C3, C4, C5) N antigens, based on comorbidity. Cognitive impairement (row 1); Cardiovascular disease (row 2); History of cancer (row 3); Diabetes (row 4); Chronic lung disease (row 5). RLU: relative light units.

**Supplemental Figure 4.** Antibody responses based on age stratified by previous infection status

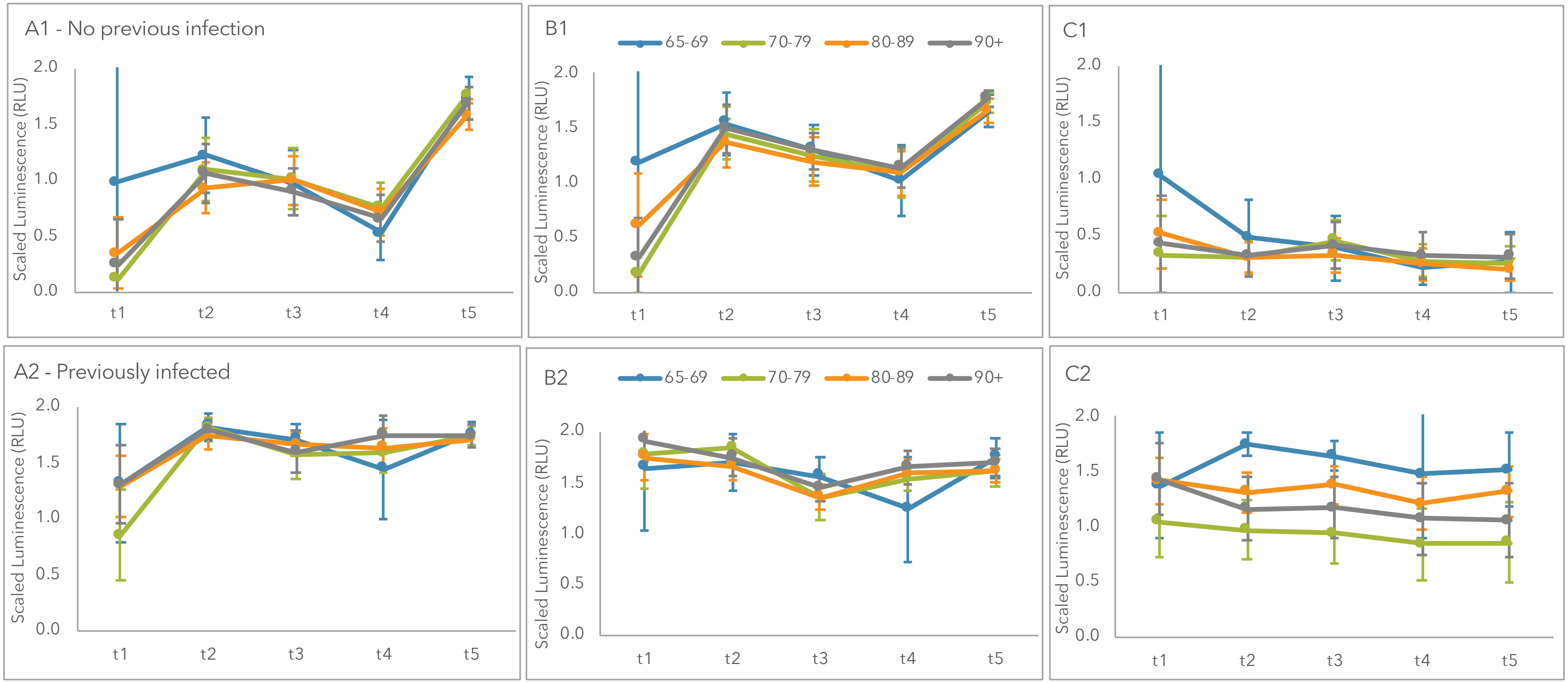

Antibody responses to (A1, A2) RBD, (B1, B2) S and (C1, C2) N antigens, based on age group, stratified by absence (top row) or presence (bottom row) of previous SARS-CoV2 infection status. RLU: relative light units.

**Supplemental Figure 5.** Antibody responses based on age stratified by homologous vs. heterologous use of mRNA vaccines

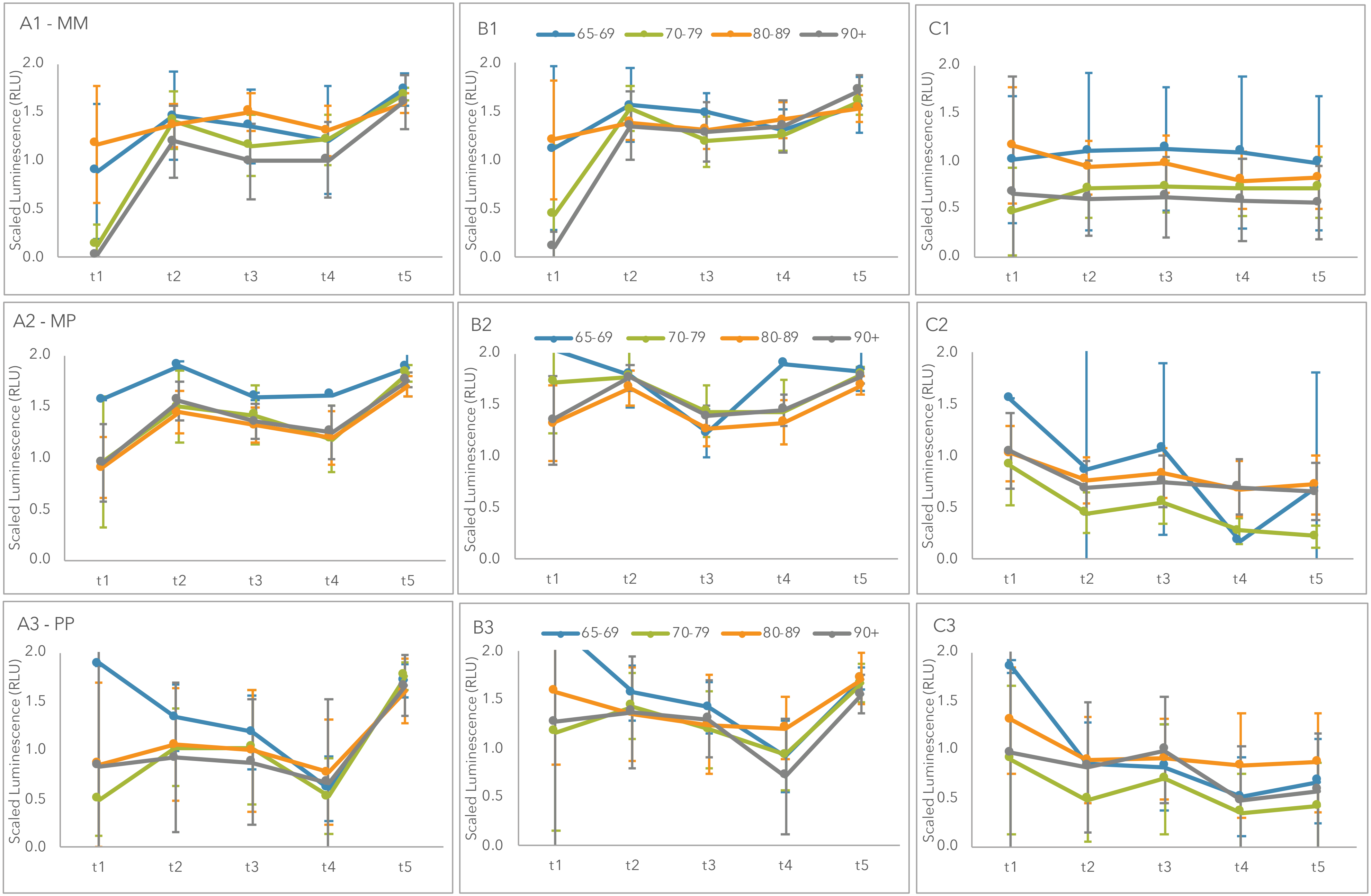

Antibody responses to (A1, A2, A3) RBD, (B1, B2, B3) S and (C1, C2, C3) N antigens, based on age group, stratified by homologous vs. heterologous use of mRNA vaccines. M: Moderna (mRNA-1273). P: Pfizer (BNT162b2). RLU: relative light units.

**Supplemental Figure 6.** Antibody responses based on sex stratified by previous infection status

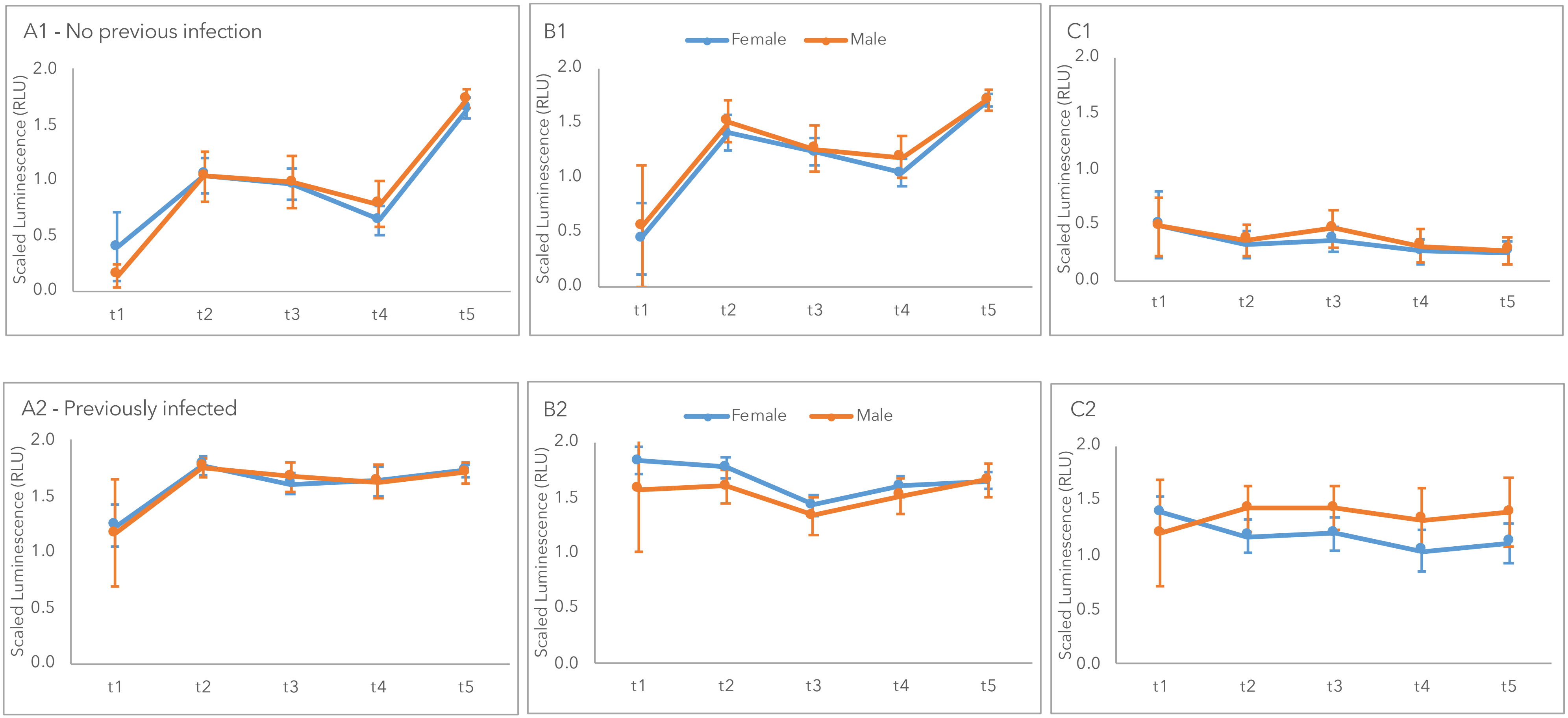

Antibody responses to (A1, A2) RBD, (B1, B2) S and (C1, C2) N antigens, based on sex, stratified by absence (top row) or presence (bottom row) of previous SARS-CoV2 infection status. RLU: relative light units.

**Supplemental Figure 7.** Antibody responses based on sex stratified by homologous vs. heterologous use of mRNA vaccines

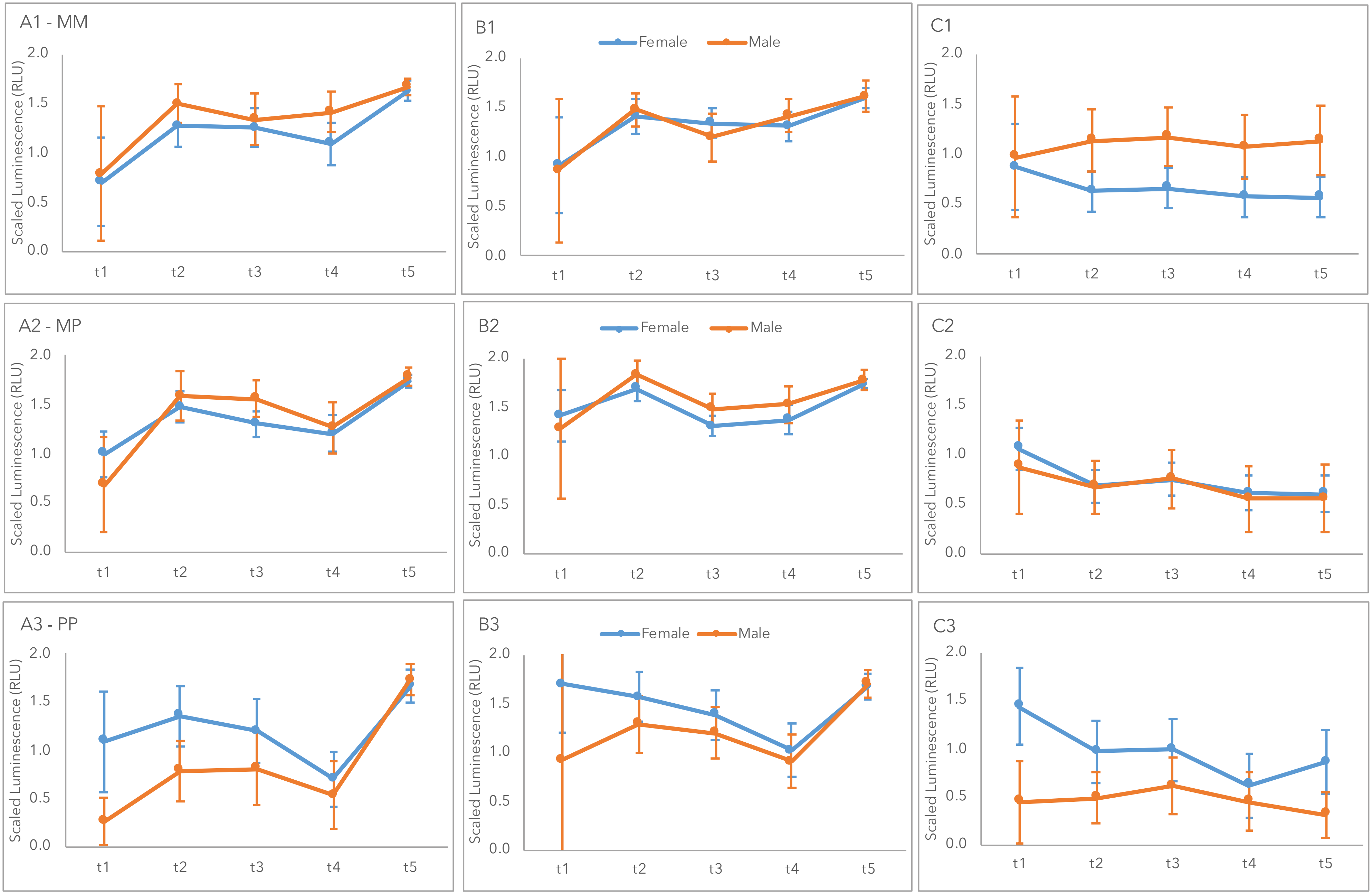

Antibody responses to (A1, A2, A3) RBD, (B1, B2, B3) S and (C1, C2, C3) N antigens, based on sex, stratified by homologous vs. heterologous use of mRNA vaccines. M: Moderna (mRNA-1273). P: Pfizer (BNT162b2). RLU: relative light units.
